# Geospatial Analysis of Individual and Community-Level Socioeconomic Factors Impacting SARS-CoV-2 Prevalence and Outcomes

**DOI:** 10.1101/2020.09.30.20201830

**Authors:** Sara J. Cromer, Chirag M. Lakhani, Deborah J Wexler, Sherri-Ann M. Burnett-Bowie, Miriam Udler, Chirag J. Patel

## Abstract

**Background:** The SARS-CoV-2 pandemic has disproportionately affected racial and ethnic minority communities across the United States. We sought to disentangle individual and census tract-level sociodemographic and economic factors associated with these disparities.

**Methods and Findings:** All adults tested for SARS-CoV-2 between February 1 and June 21, 2020 were geocoded to a census tract based on their address; hospital employees and individuals with invalid addresses were excluded. Individual (age, sex, race/ethnicity, preferred language, insurance) and census tract-level (demographics, insurance, income, education, employment, occupation, household crowding and occupancy, built home environment, and transportation) variables were analyzed using linear mixed models predicting infection, hospitalization, and death from SARS-CoV-2.

Among 57,865 individuals, per capita testing rates, individual (older age, male sex, non-White race, non-English preferred language, and non-private insurance), and census tract-level (increased population density, higher household occupancy, and lower education) measures were associated with likelihood of infection. Among those infected, individual age, sex, race, language, and insurance, and census tract-level measures of lower education, more multi-family homes, and extreme household crowding were associated with increased likelihood of hospitalization, while higher per capita testing rates were associated with decreased likelihood. Only individual-level variables (older age, male sex, Medicare insurance) were associated with increased mortality among those hospitalized.

**Conclusions:** This study of the first wave of the SARS-CoV-2 pandemic in a major U.S. city presents the cascade of outcomes following SARS-CoV-2 infection within a large, multi-ethnic cohort. SARS-CoV-2 infection and hospitalization rates, but not death rates among those hospitalized, are related to census tract-level socioeconomic characteristics including lower educational attainment and higher household crowding and occupancy, but not neighborhood measures of race, independent of individual factors.

## INTRODUCTION

As infections with SARS-CoV-2 and its associated illness, COVID-19, have surged across the world, multiple studies have identified higher rates of infection, hospitalization, and mortality among minority populations.^1–12^ Disparities in infection rates exist even among healthcare workers^13^ and children,^14,15^ with disparities in mortality rates greatest among young individuals^16^ and growing as the pandemic spreads.^17^ To date, the relative contributions of pathophysiologic and socioeconomic factors contributing to these disparities remains unclear.

Many studies demonstrate that racial and ethnic minority communities in the U.S. have higher rates of chronic disease,^18,19^ which likely contributes to the increased risk of severe illness and death related to SARS-CoV-2 infection seen in these communities. However, social determinants of health likely contribute both to this underlying disparity in chronic disease prevalence, with some claiming that socioeconomic and environmental factors may play an even stronger role in disease than individual factors,^20^ and to risk for SARS-CoV-2. This report aims to disentangle the relative contributions of individual and neighborhood sociodemographic and economic factors associated with individual risk.

Many experts in the field of health disparities have authored viewpoints suggesting factors which may contribute to risk of infection with or adverse outcomes from SARS-CoV-2, including language barriers, lower health literacy, higher population density, household overcrowding, reliance on public transit, overrepresentation among essential workers, lack of paid sick leave, inability to work from home, and limited physical and financial access to healthcare.^18,21–26^ However, despite widespread interest in this topic,^3–6^ relatively few studies have reported data examining robust socioeconomic concepts in relationship to SARS-CoV-2 outcomes. Many COVID-19 surveillance studies on this topic do not present individual data, but aggregated outcomes over large and diverse geographic areas (e.g. counties). Those studies which do present individual outcome data have included variable measures of socioeconomic status, and many of these studies are limited by small sample sizes, ethnically homogeneous populations, or examination of only a single socioeconomic measure (e.g. income) or SARS-CoV-2-related outcome (e.g. hospitalization, without consideration of the upstream outcome of infection).

Further, many studies capture area-level socioeconomic measures on a county, city/town, or zip code level, which obscures the granular variation in socioeconomic factors over adjacent areas. Census tracts represent geographical units that are smaller in population size than zip codes (average population 4,000 vs 30,000) and represent more homogeneous socioeconomic regions,^27^ leading to stronger associations with health outcomes.^28^ We hypothesized that high resolution geocoding to the census tract-level would capture more variation in COVID-19 cases and disease trajectory related to sociodemographic and economic factors.

In this study, we examine both individual and census tract-level indicators to identify factors associated with SARS-CoV-2 infection, hospitalization among infected individuals, and death among hospitalized individuals in a large and diverse cohort seeking care within an integrated healthcare system in New England.

## METHODS

### DATA SOURCES

Individual demographics, address, laboratory values, diagnoses, hospitalizations, and deaths were obtained from the electronic medical record (EMR) of Mass General Brigham (MGB, formerly Partners HealthCare), a large, integrated healthcare system serving Eastern Massachusetts. Census tract sociodemographic, economic, and built environment characteristics were obtained from the 2014-2018 American Community Survey (ACS).

### STUDY POPULATION

All individuals who received viral polymerase chain reaction (PCR) testing for SARS-CoV-2 in the MGB system between February 1, 2020 and June 21, 2020, a window beginning approximately two months before and ending three months after the peak of the Massachusetts surge, were included. Individual addresses were geocoded to a latitude and longitude (using the DeGAUSS geocoder,^29^ Supplemental Figure 1) from which census tracts were assigned. Individuals were excluded if their address was invalid, unlisted, a PO box number, or located outside Massachusetts; they did not have a valid test result (e.g. test inconclusive); their age was < 18 years; or they were employed by the MGB system (due to different exposures and thresholds for testing compared to the general population).

### STUDY OUTCOMES AND COVARIATES

Outcomes included (1) infection with SARS-CoV-2, (2) hospitalization related to SARS-CoV-2 among those testing positive, and (3) death among those hospitalized (see Figure 1 for population included in each outcomes analysis). SARS-CoV-2 infection was defined by positive PCR testing at an MGB facility. Hospitalizations were considered related to SARS-CoV-2 if a positive test occurred during or up to ten days prior to the inpatient admission or if a positive test occurred outside this window but inpatient encounter ICD10 codes suggested a SARS-CoV-2-related admission (B97.29, Z03.818, or Z20.828, recommended by the CDC prior to the availability of codes specific for SARS-CoV-2;^30^ or U07.1, “COVID-19,” which became effective as of April 1, 2020). Any death following a related hospitalization was considered SARS-CoV-2-related.

**Figure 1:**
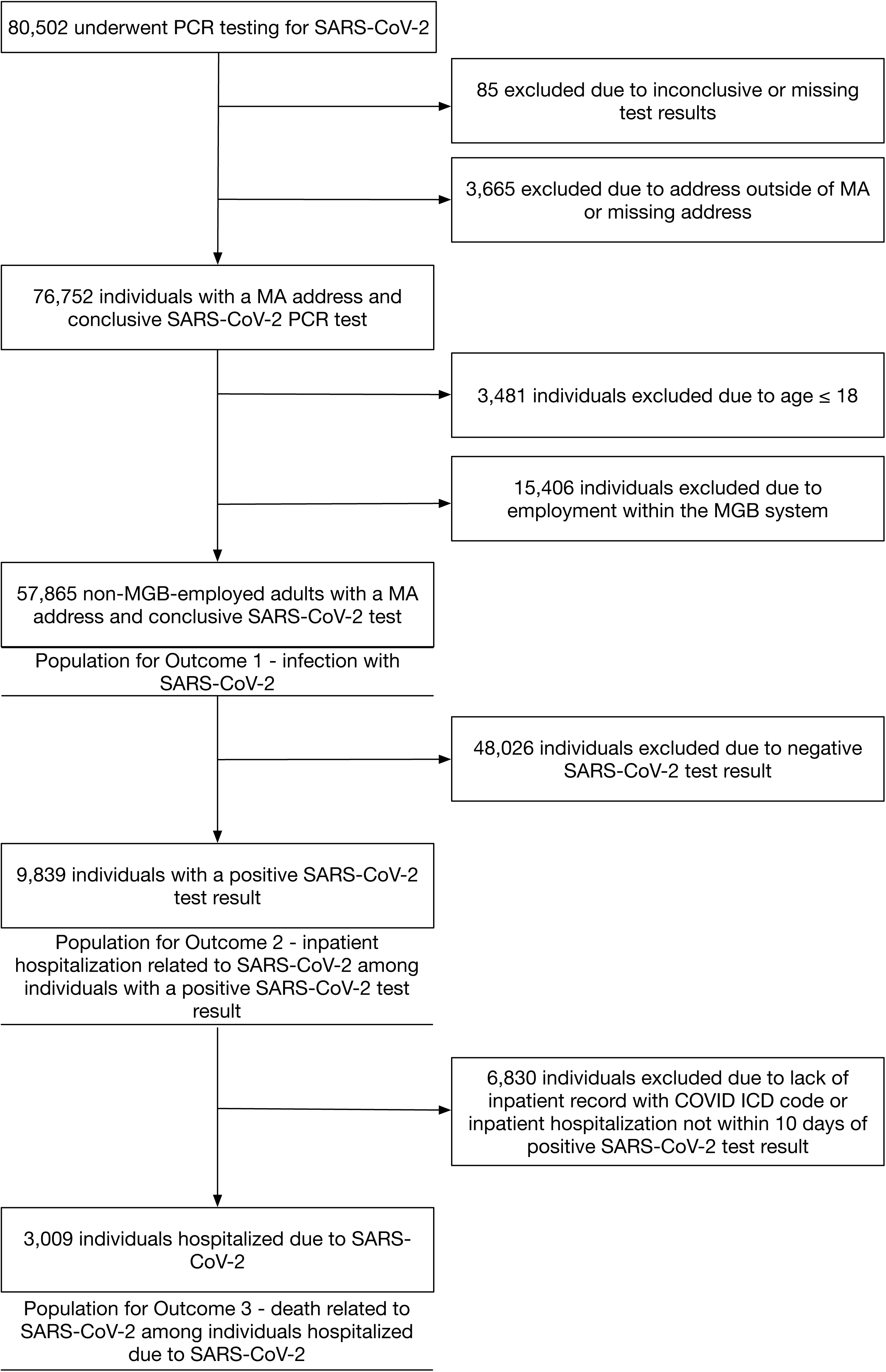
Flow diagram of individuals included in analyses of SARS-CoV-2 infection, hospitalization, and death.

Individual demographic variables included age, sex, race/ethnicity, and preferred language, each obtained by self-report upon registration in the hospital system. Racial and ethnic variables were collapsed into non-Hispanic White, non-Hispanic Black, non-Hispanic Asian, Hispanic, and Other categories, and preferred languages were categorized as English, Spanish, or Other. In patients with self-reported “Other” or missing race, self-reported country of origin and language were used to infer race when available (n=859 of 4,363; Supplemental Table 1-2). A sensitivity analysis was performed in which only those with missing, and not of “other,” race were reassigned based on either country of origin or preferred language. Insurance was categorized as private, non-Medicare (including private health insurance without any Medicare plans), Medicare (including any Medicare plan among listed insurances), public (including Medicaid, Mass Health, and other public insurance plans excluding Medicare), and safety net/uninsured (including those with safety net, self-pay, or no insurance listed).

Census tract-level variables included population density and measures of demographics, health insurance, income, education, employment, occupation, household crowding (individuals per room) and occupancy (individuals per household), built home environment (housing units per building), and transportation use, all reported as continuous percentages in ACS data tables (Supplemental Table 3). ACS variables with low variability in our sample (defined as ≥ 75% of census tracts having the same value) were dichotomized (Supplemental Table 4). Essential workers were classified by applying the U.S. Department of Homeland Security “Advisory Memorandum on Identification of Essential Critical Infrastructure Workers During COVID-19 Response” to ACS Table C24050. Per capita testing rates were calculated as the number of individuals tested in the MGB system living in that census tract divided by the total population of the tract. SARS-CoV-2 positivity rates are reported as the total number of positive individuals divided by the total number of tested individuals in each census tract.

### STATISTICAL ANALYSIS

Baseline characteristics are reported using means and standard deviations for continuous variables and proportions for categorical variables.

All models were fit using a logistic mixed model with a “matern kernel” as a random effect to account for spatial autocorrelation, using latitude and longitude of the address to estimate spatial similarity.^31^ The matern kernel has multiple hyperparameters. For each of the three outcomes (Figure 2a) we used the “base variables” (including all individual-level characteristics and census tract per capita testing rates; Table 1, Figure 2b) to perform a “grid search” to find the range and knot parameters that gave the best model (Figure 2c; the “base models”). We used the same parameters in subsequent analyses. We then fit logistic mixed models using all base variables plus each census tract variable alone (the “base-plus models”). We accounted for multiple hypothesis testing for census tract-level variables in the base-plus models using the false discovery rate (FDR) adjustment procedure where an FDR-adjusted p-value of < 0.05 was deemed significant (Table 2). Finally, we fit multivariable logistic mixed models with all base variables and all census tract variables which fell under the FDR significance threshold in the base-plus models (the “full models”). For any cluster of similar census variables (e.g. percent of households with ≥ 5 people, percent of households with ≥ 6 people) we selected the variable with the lowest p-value to be included in the full model. We exponentiated all coefficients in order to report odds ratio per 1-unit change of each variable (percentage point, year in age, or versus the referent [Tables 1-2]). In the full multivariable model, we assessed significance and independent association using a p-value < 0.05. All analyses were performed using R version 3.6.2.

**Table 1:** Baseline Characteristics of Individuals Included in the Analysis for Each Outcome, Demonstrating the Cascade of COVID-19 Care.

|  | Total Tested<sup>a</sup><br>(n=57,865) | Any Positive Test<sup>b</sup><br>(n=9,839, 17.0%) | Hospitalized, given positive test<sup>b</sup><br>(n=3,009, 30.6%) | Deceased, given hospitalization<sup>b</sup><br>(n=524, 17.4%) |
| --- | --- | --- | --- | --- |
| Age, mean (SD) | 52.32 (19.31) | 51.23 (19.89) | 62.7 (18.90) | 77.05 (13.42) |
| Age Groups, n (%) |  |  |  |  |
| 18-49 years | 25,877 (44.72) | 4,827 (18.65) | 752 (15.58) | 19 (2.53) |
| 50-64 years | 15,041 (25.99) | 2,475 (16.46) | 784 (31.68) | 73 (9.31) |
| 65-79 years | 11,837 (20.46) | 1,479 (12.49) | 813 (54.97) | 176 (21.65) |
| ≥ 80 years | 5,110 (8.83) | 1,058 (20.70) | 660 (62.38) | 256 (38.79) |
| Sex, n (%) |  |  |  |  |
| Female | 32,260 (55.75) | 4,966 (15.39) | 1,396 (28.11) | 224 (16.05) |
| Male | 25,586 (44.22) | 4,871 (19.04) | 1,613 (33.11) | 300 (18.60) |
| Race/Ethnicity, n (%)<sup>c</sup> |  |  |  |  |
| White, non-Hispanic | 34,398 (59.45) | 3,235 (9.40) | 1,307 (40.40) | 320 (24.48) |
| Black, non-Hispanic | 5,528 (9.55) | 1,239 (22.41) | 468 (37.77) | 72 (15.38) |
| Asian, non-Hispanic | 2,077 (3.59) | 298 (14.35) | 119 (39.93) | 16 (13.45) |
| Hispanic | 12,358 (21.36) | 4,257 (34.45) | 985 (23.14) | 94 (9.54) |
| Other | 649 (1.12) | 112 (17.26) | 39 (34.82) | 4 (10.26) |
| Missing | 2,855 (4.93) | 698 (24.45) | 91 (13.04) | 18 (19.78) |
| Language, n(%) |  |  |  |  |
| English | 4,6086 (79.64) | 5,403 (11.72) | 1,855 (34.33) | 369 (19.89) |
| Spanish | 7,602 (13.14) | 3,289 (43.26) | 824 (25.05) | 77 (9.34) |
| Other | 2,118 (3.66) | 591 (27.90) | 279 (47.21) | 69 (24.73) |
| Missing | 2,059 (3.56) | 556 (27.00) | 51 (9.17) | 9 (17.65) |
| Insurance, n(%) |  |  |  |  |
| Private, non-Medicare | 29,740 (51.40) | 4,031 (13.55) | 935 (23.20) | 85 (9.09) |
| Medicare | 16,480 (28.48) | 2,311 (14.02) | 1,365 (59.07) | 400 (29.30) |
| Medicaid, MassHealth | 8,397 (14.51) | 2,435 (29.00) | 630 (25.87) | 36 (5.71) |
| Safety Net/Unlisted | 3,248 (5.61) | 1,062 (32.70) | 79 (7.44) | 3 (3.80) |

<sup>a</sup> Percentages represent the percent of individuals tested who fell into a category. (e.g. 44.22% of all individuals tested were male).
<sup>b</sup> Percentages represent the percent of individuals experiencing that outcome, among all eligible to experience the outcome in that cell (e.g. 19.04% of men tested for SARS-CoV-2 received a positive test result).
<sup>c</sup> Includes reassigned race (based on country of origin or language) for those with recorded Other or Missing race.

**Table 2.** Individual and Census Tract Correlates of Individual SARS-CoV-2 Test Positivity, SARS-CoV-2 Related Admission, and SARS-CoV-2-related Death in Base (Individual Characteristics only), Base-Plus (Base Model Covariates plus Single Census Tract-Level Variables), and Final Multivariable Modelsxs

|  | SARS-CoV-2 Infection |  |  | SARS-CoV-2-related Hospitalization |  |  | SARS-CoV-2-related Mortality |  |  |
| --- | --- | --- | --- | --- | --- | --- | --- | --- | --- |
|  | Base Model<sup>a</sup> | Base-Plus Models<sup>b</sup> | Full Model<sup>c</sup> | Base Model<sup>a</sup> | Base-Plus Models<sup>b</sup> | Full Model<sup>c</sup> | Base Model<sup>a</sup> | Base-Plus Models<sup>b</sup> | Full Model<sup>c</sup> |
| Number of individuals in analysis, n |  | 57,865 |  |  | 9,839 |  |  | 3,009 |  |
| **Individual Characteristics** |  |  |  |  |  |  |  |  |  |
| Age | **1.01 (1.01, 1.01)** |  | **1.01 (1.01, 1.01)** | **1.04 (1.04, 1.04)** |  | **1.04 (1.04, 1.04)** | **1.06 (1.05, 1.07)** |  | **1.06 (1.05, 1.07)** |
| Sex |  |  |  |  |  |  |  |  |  |
| Female | Ref |  | Ref | Ref |  | Ref | Ref |  | Ref |
| Male | **1.30 (1.24, 1.37)** |  | **1.30 (1.24, 1.37)** | **1.50 (1.36, 1.66)** |  | **1.50 (1.36, 1.66)** | **1.64 (1.32, 2.04)** |  | **1.64 (1.32, 2.04)** |
| Race/Ethnicity<sup>d</sup> |  |  |  |  |  |  |  |  |  |
| White, non-Hispanic | Ref |  | Ref | Ref |  | Ref | Ref |  | Ref |
| Black, non-Hispanic | **2.72 (2.50, 2.95)** |  | **2.62 (2.41, 2.86)** | **1.47 (1.23, 1.74)** |  | **1.39 (1.16, 1.65)** | 0.81 (0.57, 1.14) |  | 0.81 (0.57, 1.14) |
| Asian, non-Hispanic | **1.37 (1.19, 1.58)** | Base | **1.36 (1.18, 1.56)** | **1.57 (1.17, 2.11)** | Base | **1.53 (1.14, 2.06)** | 0.58 (0.30, 1.11) | Base | 0.58 (0.30, 1.11) |
| Hispanic | **2.26 (2.07, 2.46)** | Model | **2.20 (2.02, 2.40)** | **0.81 (0.66, 0.99)** | Model | **0.77 (0.63, 0.94)** | 1.38 (0.80, 2.36) | Model | 1.38 (0.80, 2.36) |
| Other | **1.84 (1.49, 2.28)** | Plus | **1.82 (1.47, 2.25)** | **1.59 (1.01, 2.52)** | Plus | 1.53 (0.97, 2.42) | 0.43 (0.14, 1.34) | Plus | 0.43 (0.14, 1.34) |
| Missing | **2.17 (1.88, 2.49)** | Single | **2.14 (1.86, 2.46)** | **0.70 (0.50, 0.97)** | Single | **0.68 (0.49, 0.95)** | 1.05 (0.55, 2.00) | Single | 1.05 (0.55, 2.00) |
| Language, n(%) |  | Census |  |  | Census |  |  | Census |  |
| English | Ref | Tract-Level Variables | Ref | Ref | Tract-Level Variables | Ref | Ref | Tract-Level Variables | Ref |
| Spanish | **2.37 (2.18, 2.58)** |  | **2.35 (2.15, 2.56)** | **1.27 (1.05, 1.54)** |  | **1.25 (1.03, 1.52)** | 0.62 (0.35, 1.09) |  | 0.62 (0.35, 1.09) |
| Other | **1.94 (1.74, 2.16)** | Below | **1.91 (1.71, 2.13)** | 1.22 (0.98, 1.51) | Below | 1.19 (0.96, 1.47) | 1.26 (0.87, 1.84) | Below | 1.26 (0.87, 1.84) |
| Missing | **1.64 (1.41, 1.92)** |  | **1.63 (1.40, 1.91)** | **0.43 (0.29, 0.64)** |  | **0.42 (0.28, 0.63)** | 1.17 (0.48, 2.85) |  | 1.17 (0.48, 2.85) |
|  | Base Model<sup>a</sup> | Base-Plus Models<sup>b</sup> | Full Model<sup>c</sup> | Base Model<sup>a</sup> | Base-Plus Models<sup>b</sup> | Full Model<sup>c</sup> | Base Model<sup>a</sup> | Base-Plus Models<sup>b</sup> | Full Model<sup>c</sup> |
| Insurance, n(%) |  |  |  |  |  |  |  |  |  |
| Private, non-Medicare | Ref |  | Ref | Ref |  | Ref | Ref |  | Ref |
| Medicare | 0.95 (0.88, 1.02) |  | 0.94 (0.88, 1.01) | **2.22 (1.93, 2.56)** |  | **2.20 (1.92, 2.54)** | **1.87 (1.41, 2.49)** |  | **1.87 (1.41, 2.49)** |
| Medicaid, MassHealth | **1.21 (1.13, 1.30)** |  | **1.21 (1.13, 1.29)** | **1.59 (1.38, 1.82)** |  | **1.57 (1.37, 1.81)** | 1.02 (0.65, 1.58) |  | 1.02 (0.65, 1.58) |
| Safety Net/Unlisted | **1.25 (1.14, 1.37)** |  | **1.24 (1.13, 1.37)** | **0.36 (0.28, 0.47)** |  | **0.36 (0.28, 0.47)** | 0.71 (0.21, 2.39) |  | 0.71 (0.21, 2.39) |
| **Census Tract Characteristics** |  |  |  |  |  |  |  |  |  |
| Per Capita Testing Rate | **2262.11 (259.09, 19750.08)** |  | **1288.28 (125.72, 13201.02)** | **0.00 (0.00, 0.29)** |  | **0.00 (0.00, 0.03)** | 0.39 (0.00, 296.76) |  | 0.39 (0.00, 296.76) |
| Percent age ≥ 65 |  | 0.78 (0.43, 1.44) |  |  | 0.63 (0.18, 2.23) |  |  | 0.82 (0.12, 5.81) |  |
| Race/Ethnicity, mean (IQR) |  |  |  |  |  |  |  |  |  |
| % White, non-Hispanic |  | **0.60 (0.49, 0.74)** |  |  | 0.73 (0.48, 1.12) |  |  | 0.55 (0.31, 1.00) |  |
| % Black, non-Hispanic |  | **2.21 (1.51, 3.23)** | 1.53 (0.96, 2.46) |  | 0.68 (0.32, 1.43) |  |  | 1.51 (0.58, 3.92) |  |
| % Asian, non-Hispanic |  | 0.95 (0.58, 1.58) | 1.11 (0.69, 1.79) |  | 1.19 (0.42, 3.37) |  |  | 1.46 (0.26, 8.27) |  |
| % Hispanic |  | **1.67 (1.22, 2.29)** | 1.30 (0.83, 2.04) |  | **2.29 (1.23, 4.26)** | 0.73 (0.31, 1.75) |  | 1.98 (0.78, 5.01) |  |
| % Foreign-born |  | **1.99 (1.41, 2.81)** | 0.80 (0.36, 1.76) |  | **3.47 (1.74, 6.92)** | 1.43 (0.58, 3.48) |  | 1.44 (0.49, 4.23) |  |
| Insurance |  |  |  |  |  |  |  |  |  |
| % Private, non-Medicare |  | **0.66 (0.51, 0.85)** | 0.80 (0.36, 1.76) |  | 0.53 (0.32, 0.88) |  |  | 0.51 (0.23, 1.15) |  |
| % Medicare |  | 0.68 (0.34, 1.38) |  |  | 0.96 (0.22, 4.18) |  |  | 0.59 (0.05, 6.45) |  |
| % Medicaid, MassHealth |  | **1.50 (1.14, 1.98)** | 0.50 (0.22, 1.15) |  | 1.58 (0.90, 2.76) |  |  | 1.75 (0.71, 4.29) |  |
| % Safety Net/None |  | 33.50 (0.00, 2.20E7) |  |  | 1438.03 (0.00, 3.59E15) |  |  | 599647228.33 (0.00, 1.15E29) |  |
| % Safety Net/None, dichotomized |  | 0.99 (0.92, 1.06) |  |  | 1.01 (0.88, 1.17) |  |  | 1.10 (0.82, 1.47) |  |
|  | Base Model<sup>a</sup> | Base-Plus Models<sup>b</sup> | Full Model<sup>c</sup> | Base Model<sup>a</sup> | Base-Plus Models<sup>b</sup> | Full Model<sup>c</sup> | Base Model<sup>a</sup> | Base-Plus Models<sup>b</sup> | Full Model<sup>c</sup> |
| Population and Population Density |  |  |  |  |  |  |  |  |  |
| Log (Total Population) |  | 0.90 (0.74, 1.09) |  |  | 0.90 (0.60, 1.34) |  |  | 1.74 (0.81, 3.73) |  |
| Log (Population Density) |  | **1.24 (1.14, 1.36)** | **1.14 (1.03, 1.27)** |  | **1.25 (1.04, 1.51)** | 0.99 (0.80, 1.24) |  | 1.14 (0.86, 1.51) |  |
| Income and Income Inequality |  |  |  |  |  |  |  |  |  |
| Median household income |  | 0.44 (0.14, 1.37) |  |  | **0.04 (0.00, 0.38)** | 1.29 (0.02, 107.59) |  | 0.05 (0.00, 1.33) |  |
| % public assistance income |  | 2.06 (0.67, 6.31) |  |  | 0.34 (0.03, 3.42) |  |  | 1.38 (0.01, 154.02) |  |
| GINI Index |  | 0.96 (0.56, 1.66) |  |  | 0.57 (0.18, 1.81) |  |  | 1.22 (0.16, 9.45) |  |
| Poverty |  |  |  |  |  |  |  |  |  |
| % living below 100% federal poverty level |  | 1.13 (0.75, 1.69) |  |  | 1.70 (0.75, 3.86) |  |  | 1.59 (0.40, 6.31) |  |
| % living below 150% federal poverty level |  | 1.40 (1.03, 1.91) |  |  | 1.76 (0.94, 3.31) |  |  | 1.81 (0.64, 5.16) |  |
| % living below 200% federal poverty level |  | 1.29 (0.99, 1.69) |  |  | **2.07 (1.20, 3.55)** | 0.63 (0.26, 1.54) |  | 1.85 (0.76, 4.51) |  |
| Education |  |  |  |  |  |  |  |  |  |
| % without high school diploma |  | **2.64 (1.69, 4.14)** | 1.39 (0.67, 2.91) |  | **6.28 (2.55, 15.49)** | **5.19 (1.15, 23.50)** |  | 1.82 (0.41, 8.03) |  |
| % with college degree |  | **0.60 (0.47, 0.76)** | **0.63 (0.40, 0.99)** |  | **0.52 (0.33, 0.83)** | 0.84 (0.36, 1.92) |  | 0.60 (0.32, 1.13) |  |
| Employment and Occupation |  |  |  |  |  |  |  |  |  |
| % unemployed |  | **7.58 (1.78, 32.32)** | 2.82 (0.59, 13.52) |  | 10.35 (0.50, 215.08) |  |  | 2.88 (0.01, 965.45) |  |
| % essential workers |  | **1.56 (1.15, 2.12)** | 0.70 (0.43, 1.13) |  | 2.19 (1.20, 4.02) |  |  | 1.45 (0.59, 3.58) |  |
|  | Base Model<sup>a</sup> | Base-Plus Models<sup>b</sup> | Full Model<sup>c</sup> | Base Model<sup>a</sup> | Base-Plus Models<sup>b</sup> | Full Model<sup>c</sup> | Base Model<sup>a</sup> | Base-Plus Models<sup>b</sup> | Full Model<sup>c</sup> |
| Household Crowding |  |  |  |  |  |  |  |  |  |
| % with >1 occupant per room |  | 2.95 (1.08, 8.08) |  |  | 8.89 (1.18, 67.10) |  |  | 12.65 (0.25, 632.77) |  |
| % with >1.5 occupants per room |  | 2.91 (0.32, 26.83) |  |  | 6.88 (0.08, 578.34) |  |  | 157.80 (0.02, 1628070.04) |  |
| % with >2 occupants per room |  | 2.49 (0.07, 84.40) |  |  | 11980.64 (8.68, 1.65E7) |  |  | 0.11 (0.00, 2.05E6) |  |
| % with >2 occupants per room, dichotomized |  | 1.00 (0.94, 1.07) |  |  | **1.16 (1.02, 1.32)** | **1.14 (1.01, 1.30)** |  | 0.95 (0.73, 1.24) |  |
| Household Occupancy |  |  |  |  |  |  |  |  |  |
| % households with ≥ 2 members |  | 1.24 (0.90, 1.71) |  |  | 0.73 (0.38, 1.40) |  |  | 0.85 (0.28, 2.59) |  |
| % households with ≥ 3 members |  | **1.54 (1.15, 2.05)** |  |  | 0.95 (0.53, 1.70) |  |  | 0.81 (0.28, 2.35) |  |
| % households with ≥ 4 members |  | **1.72 (1.20, 2.47)** |  |  | 0.70 (0.33, 1.45) |  |  | 0.87 (0.23, 3.34) |  |
| % households with ≥ 5 members |  | **3.13 (1.72, 5.72)** | **2.20 (1.13, 4.30)** |  | 0.92 (0.27, 3.08) |  |  | 1.74 (0.17, 18.02) |  |
| % households with ≥ 6 members |  | 3.36 (1.12, 10.10) |  |  | 13.29 (1.53, 115.08) |  |  | 9.88 (0.15, 630.50) |  |
| % households with ≥ 7 members |  | 5.28 (0.74, 37.49) |  |  | 32.06 (0.61, 1697.21) |  |  | 8.07 (0.00, 23380.62) |  |
| Housing Factors |  |  |  |  |  |  |  |  |  |
| Median Home Value |  | 0.88 (0.71, 1.10) |  |  | 0.63 (0.41, 0.98) |  |  | 0.49 (0.28, 0.88) |  |
| % housing units lacking plumbing facilities |  | 2.65 (0.92, 7.68) |  |  | 9.04 (0.90, 90.56) |  |  | 0.61 (0.01, 50.33) |  |
|  | Base Model<sup>a</sup> | Base-Plus Models<sup>b</sup> | Full Model<sup>c</sup> | Base Model<sup>a</sup> | Base-Plus Models<sup>b</sup> | Full Model<sup>c</sup> | Base Model<sup>a</sup> | Base-Plus Models<sup>b</sup> | Full Model<sup>c</sup> |
| Housing Type |  |  |  |  |  |  |  |  |  |
| % 1 unit in structure |  | 0.91 (0.76, 1.10) |  |  | **0.52 (0.36, 0.74)** |  |  | 0.78 (0.48, 1.29) |  |
| % ≥ 2 units in structure |  | 1.12 (0.92, 1.36) |  |  | **2.08 (1.42, 3.04)** | **1.83 (1.01, 3.31)** |  | 1.31 (0.76, 2.25) |  |
| % ≥ 3 units in structure |  | 0.99 (0.82, 1.21) |  |  | 1.43 (0.98, 2.11) |  |  | 1.34 (0.77, 2.33) |  |
| % ≥ 5 units in structure |  | 0.92 (0.77, 1.11) |  |  | 0.89 (0.62, 1.30) |  |  | 0.93 (0.50, 1.73) |  |
| % ≥ 10 units in structure |  | 0.94 (0.78, 1.13) |  |  | 0.96 (0.65, 1.40) |  |  | 1.08 (0.56, 2.08) |  |
| % ≥ 20 units in structure |  | 0.90 (0.74, 1.09) |  |  | 0.83 (0.56, 1.23) |  |  | 0.96 (0.48, 1.93) |  |
| % ≥ 50 units in structure |  | 0.85 (0.68, 1.06) |  |  | 0.81 (0.51, 1.28) |  |  | 0.86 (0.38, 1.95) |  |
| Transportation |  |  |  |  |  |  |  |  |  |
| % commute by walk |  | 1.07 (0.61, 1.89) |  |  | 1.40 (0.45, 4.35) |  |  | 0.75 (0.17, 3.26) |  |
| % commute by public transit |  | 1.09 (0.73, 1.62) |  |  | 1.97 (0.91, 4.25) |  |  | 1.69 (0.61, 4.63) |  |
| % commute by vehicle |  | 1.09 (0.78, 1.53) |  |  | 0.54 (0.28, 1.03) |  |  | 0.93 (0.43, 2.02) |  |
| % work from home |  | **0.26 (0.09, 0.75)** | 0.60 (0.19, 1.90) |  | 1.06 (0.14, 8.24) |  |  | 0.04 (0.00, 1.47) |  |

<sup>a</sup> The base model is a logistic mixed model including all individual-level characteristics and the census tract-level per capita testing rate in the Mass General Brigham (MGB) hospital system. Bolded values have a false discovery rate (FDR)-adjusted p-value < 0.05.
<sup>b</sup> The base-plus models are a series of logistic mixed models, each including the covariates of the base model with the addition of a single census tract-level variable as a means of examining the effect of that variable in the absence of other, often correlated, census tract-level covariates. Bolded values have an FDR-adjustment of p-values at a level of 0.05.
<sup>c</sup> The full multivariable model is a logistic mixed model including the covariates of the base model with the addition of all census tract-level variables which met FDR-adjusted significance thresholds in the base-plus models. If several variables within a highly related group (e.g. percent of households with ≥ 5 members, percent of household with ≥ 6 members, etc.) each met the FDR-adjusted significance threshold in its respective base-plus model, the variable with the most significant p-value was selected for inclusion in the full, multivariable model.
<sup>d</sup> Includes reassigned race (based on country of origin or language) for those with recorded Other or Missing race.

**Figure 2:**
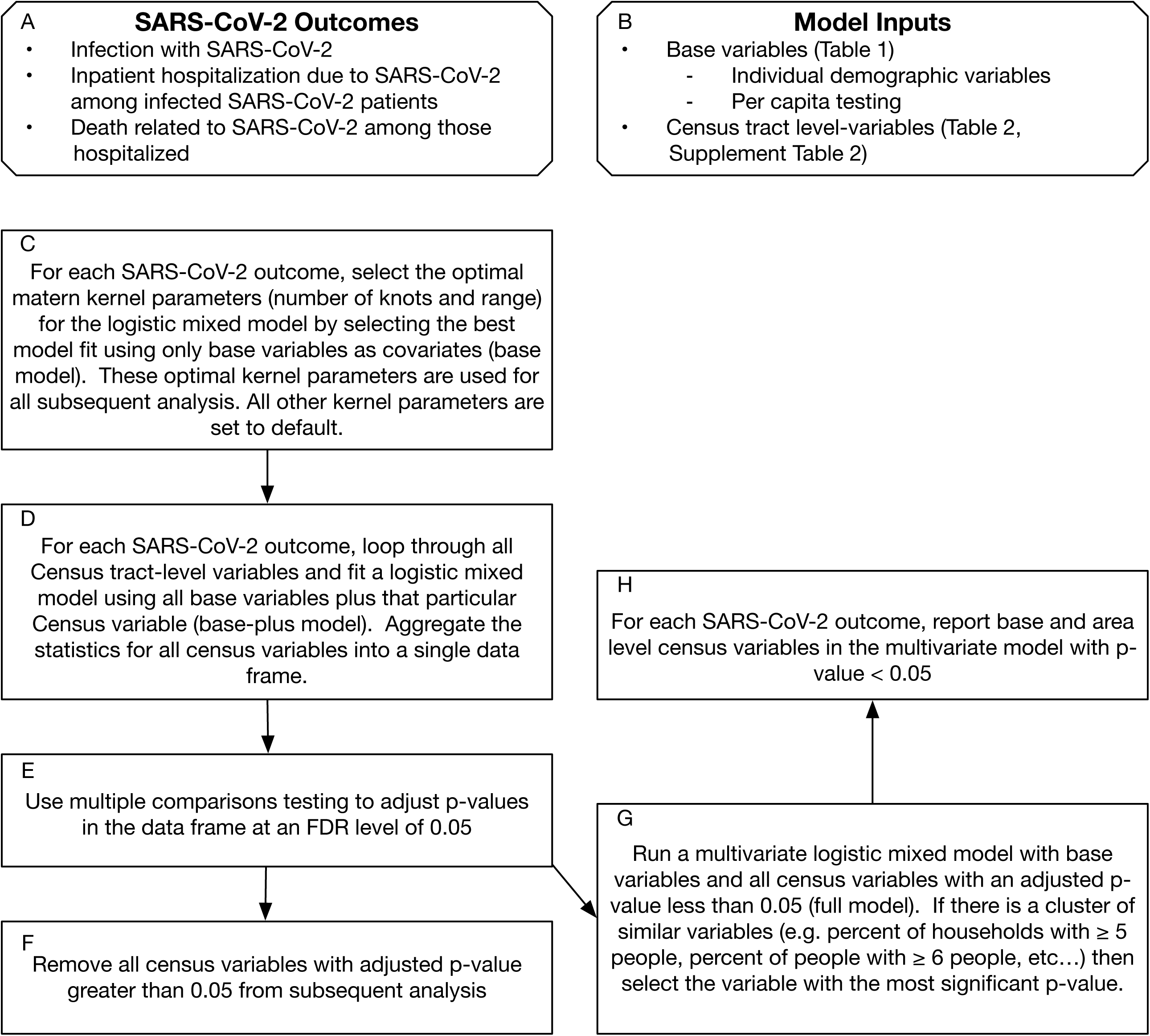
Analysis Flow Chart. FDR = False discovery rate.

This study was reviewed and deemed exempt by the Institutional Review Board of Mass General Brigham.

## RESULTS

### BASELINE CHARACTERISTICS

A total of 80,502 individuals were tested for SARS-CoV-2 in the MGB system between February 1, 2020 and June 21, 2020, and 57,865 individuals met inclusion criteria (Figure 1). Of these, 56% were female, 36% were of Asian, Black, Hispanic, or Other race/ethnicity (henceforth “non-White” for brevity), and mean age was 52.3 years. Tested individuals resided in 1,386 unique census tracts. Detailed baseline characteristics of tested individuals are presented in Table 1. Census tract characteristics in the Boston area are presented in Figure 3 and Supplemental Figure 1.

**Figure 3:**
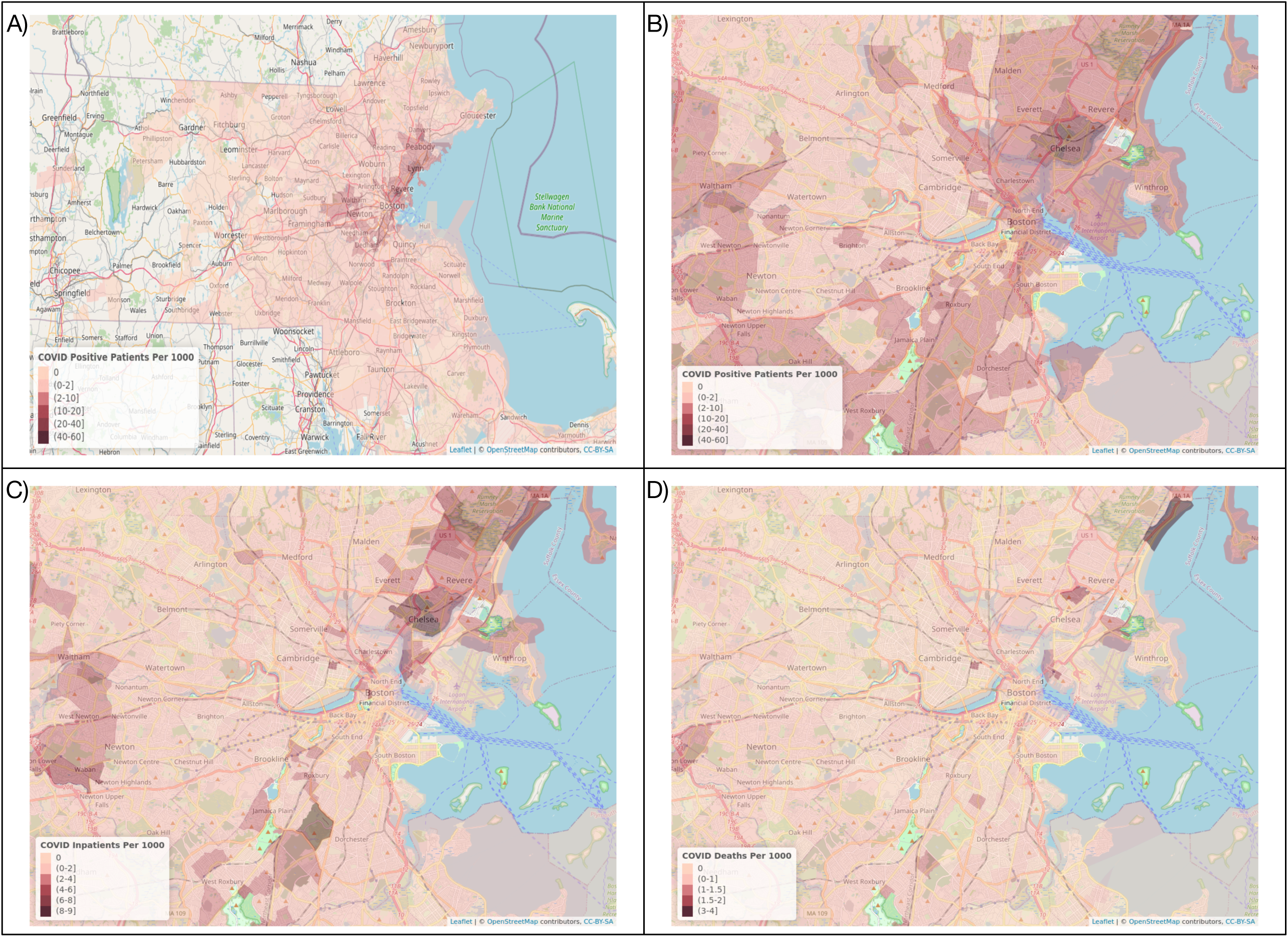
Map depicting the number of individuals per 1,000 population (A) who tested positive for SARS-CoV-2 in the MGB system in Massachusetts and (B) in the Boston area, (C) who were hospitalized at an MGB facility related to SARS-CoV-2, and (D) who died after their hospitalization. Census tracts with fewer than 5 individuals tested are excluded. Only patients tested within the MGB system are represented.

### INDIVIDUAL AND CENSUS TRACT-LEVEL FACTORS ASSOCIATED WITH SARS-CoV-2 INFECTION

Of the individuals tested for SARS-CoV-2 infection, 9,839 (17.0%) tested positive. Census tract per capita testing rates were highly associated with the likelihood of an individual testing positive. All individual-level characteristics, including older age, male sex, non-White race, preferred language being other than English, and Medicaid and uninsured/unlisted insurance status, were associated with increased likelihood of infection (Table 2). In the full multivariable models, census tract-level factors, including higher logarithm of population density (OR 1.14, 95% CI 1.03, 1.27) and increase in the percent with higher household occupancy (≥ 5 individuals per household; OR 2.20, 95% CI 1.13, 4.30) were associated with risk of infection, while increased percent with a college education (OR 0.63, 95% CI 0.40, 0.99) was associated with lower risk.

### INDIVIDUAL AND CENSUS TRACT-LEVEL FACTORS ASSOCIATED WITH HOSPITALIZATION AND DEATH RELATED TO SARS-CoV-2

Among those who tested positive, 3,009 (30.6%) required hospitalization related to SARS-CoV-2. Hospitalization was associated with individual older age; male sex; Black, Asian, or Other race; Spanish language preference; and Medicare and Medicaid insurance in all models. By contrast, higher per capita testing rates, Hispanic ethnicity, missing race or language, and uninsured/unlisted insurance status associated with lower rates of hospitalization. Census tract-level factors including presence of extreme household crowding (any homes with ≥ 2 occupants per room vs none; OR 1.14, 95% CI 1.01, 1.30), higher percent multi-family homes (OR 1.83, 95% CI 1.01, 3.31), and higher percent of individuals with less than high school education (OR 5.19, 95% CI 1.15, 23.50) were associated with hospitalization in the full model (Table 2).

Of the individuals hospitalized for SARS-CoV-2, 524 (17.4%) died. Factors associated with death during the observation period included only individual older age (OR for a 1 year increase 1.06, 95% CI 1.05, 1.07), male sex (OR 1.64, 95% CI 1.32, 2.04), and Medicare insurance (OR 1.87 vs private, 95% CI 1.41, 2.49). No census tract-level variables were associated with death in the base-plus models (Table 2). All significant results are summarized in Figure 4.

**Figure 4:**
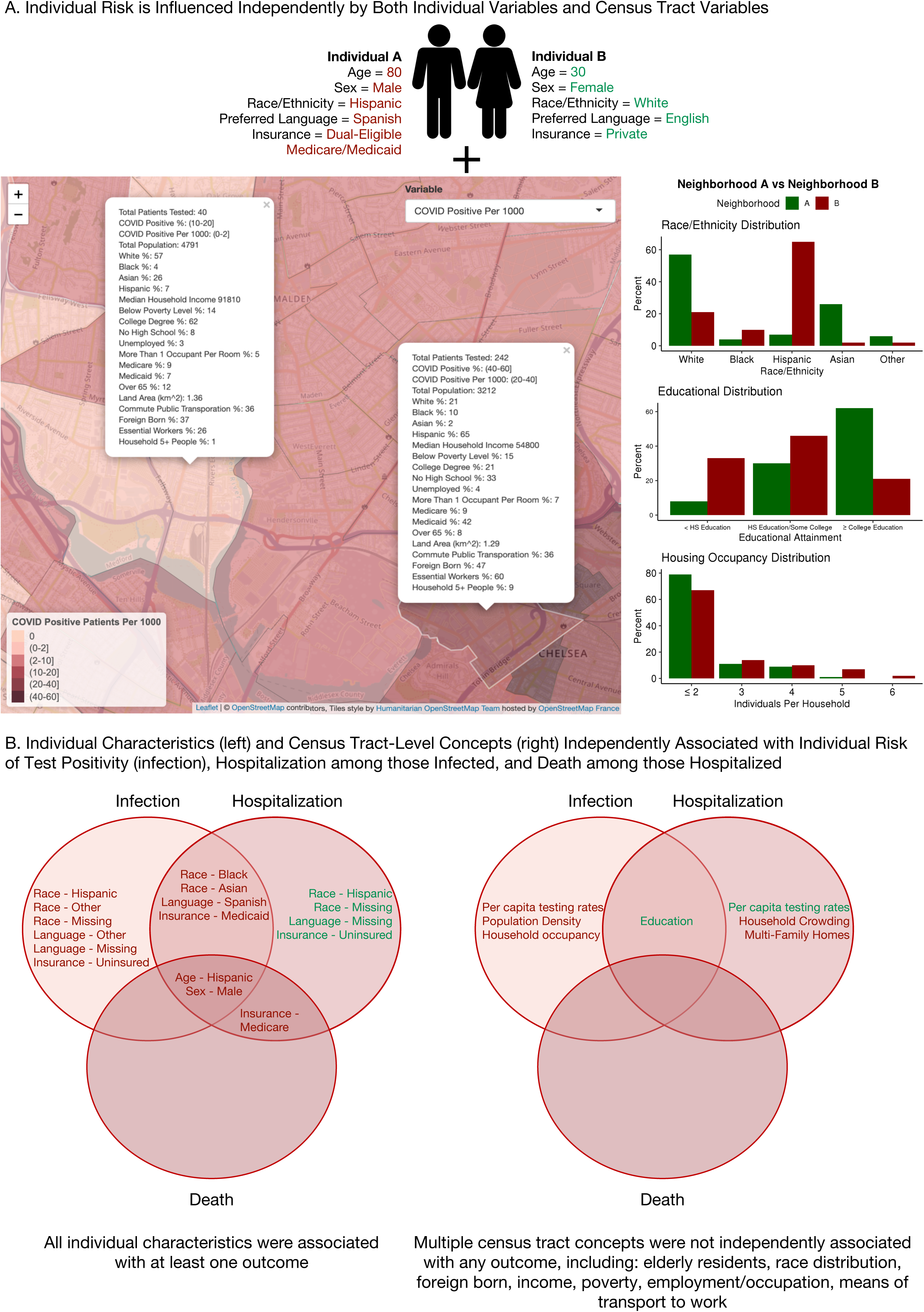
(A) Conceptual model of an individual’s aggregate risk of disease, arising from the interaction between individual and census tract-level risk factors, with red indicating a positive association with adverse outcomes (increased risk) and green indicating a negative association (decreased risk), and (B) summary of individual characteristics (left) and census tract-level concepts (right) which were independently associated with infection with SARS-CoV-2, hospitalization among those infected, and death among those hospitalized within our study sample, with red indicating a positive association with adverse outcomes (increased risk) and green indicating a negative association (decreased risk). HS = high school.

## DISCUSSION

This study demonstrates that individual SARS-CoV-2 infection in Eastern Massachusetts was associated with key census tract-level measures of higher population density, percent without a college degree, and household occupancy, independently of individual factors including older age, male sex, non-White race, non-English preferred language, and Medicaid or uninsured/unlisted insurance as compared to private insurance. Those infected with SARS-CoV-2 had higher rates of hospitalization if they lived in areas with higher rates of extreme household crowding and percent multi-family homes and lower rates of high school completion. Although census tract-level sociodemographic and economic factors were associated with infection and hospitalization, they were not associated with mortality among those hospitalized. Death was associated only with individual-level factors, including older age, male sex, and Medicare insurance (which may further capture age as a risk factor). Together, these findings suggest that while community socioeconomic factors do increase risk of infection and hospitalization, they are not clearly associated with death among those hospitalized for SARS-CoV-2, as seen in previous studies.^7,32^

Our study identified individual non-English language and census tract-level lower educational attainment as significant risk factors for both infection and hospitalization even after adjustment for race and ethnicity. To our knowledge, this has not been previously described. These findings lend support for efforts to provide easily accessible, multi-lingual educational materials targeted to all educational levels regarding social distancing practices, symptoms of SARS-CoV-2, and known risk factors for adverse outcomes. Further, we were able to refine previous associations between infection and hospitalization rates with area-level household crowding. In fact, this association is consistent with smaller studies^33,34^ and may partially explain racial disparities seen in aggregated county-level data.^19^ For example, approximately one-fifth of US residential units are unsuitable for home quarantine based on the number of rooms available per individual, with racial and ethnic minorities overrepresented among these households.^35^

By examining the full spectrum of outcomes from testing to mortality in a very large urban cohort, this study extends the developing body of research suggesting that race/ethnicity, socioeconomic status, and neighborhood factors are significantly and independently associated with risk of both infection with and adverse outcomes due to SARS-CoV-2. Multiple studies have examined aggregated larger area-level factors associated with infection or death, identifying area-level race, population density, poverty, household crowding, and lower rates of medical insurance as risk factors.^19,36,37^ By contrast, few studies have examined individual-level data to study the relationship between sociodemographic and economic factors in both SARS-CoV-2 infections and outcomes. Studies in the US and UK have identified non-White race, higher household crowding and occupancy, lower income, higher unemployment, and employment as essential or healthcare workers as correlates of infection among those tested,^15,33,38–40^ findings which are largely consistent with our analyses. However, these studies have largely examined these socioeconomic measures in isolation, without adjusting for related area-level socioeconomic measures. Similarly, non-White race, household crowding, socioeconomic deprivation indices and lower income, and Medicaid or absent health insurance have been associated with hospitalization or death,^7,8,10,34,41,42^ although it is not clear if these factors would be significant in models adjusted for other socioeconomic factors.

Our study builds on previous findings by leveraging a larger and more diverse cohort, adjusting for numerous census tract-level variables to represent diverse and correlated socioeconomic constructs, and describing the complete cascade of outcomes beginning with SARS-CoV-2 testing and including diagnosis of SARS-CoV-2, hospitalization, and death. This results in the ability to disentangle individual and neighborhood factors contributing to risk (Figure 4a). For instance, area-level measures of percent racial and ethnic minority residents, which were previously associated with SARS-CoV-2 outcomes in studies using aggregated data from our study area,^37,40^ were significant in our base-plus models (models including individual characteristics and a single census tract-level variable, e.g. percent Black residents) of infection and hospitalization; however, these variables lost significance in multivariable models adjusting for other census tract-level socioeconomic factors, such as educational attainment and household occupancy, suggesting these were the true factors associated with outcomes, rather than neighborhood racial/ethnic makeup itself. Further, we believe that analysis at the census tract-level, rather than the larger zip code or county levels, allows for improved identification of subtle associations. For example, we detect associations of census-tract educational attainment with infection rates, an association not found in a study of Massachusetts using data at the city or town-level.^40^ We find that individual age, sex, race/ethnicity, language, and insurance associate with both infection and hospitalization, while different census tract-level sociodemographic and economic factors associate with each outcome. However, even in models fully adjusted for significant community socioeconomic factors, the association of individual race/ethnicity with outcomes persists, suggesting that differences in underlying comorbidities or as yet unrecognized environmental contributors to risk may not be captured in this analysis.

One important characteristic in the discussion of disparities in SARS-CoV-2 outcomes bears further discussion. In our sample, Hispanic ethnicity and Spanish language are highly, but not completely, correlated, yet our models showed opposite directions of effect for these variables on hospitalization. To improve our understanding of these factors in isolation, we created base models for hospitalization and death, alternately excluding race/ethnicity or language (Supplemental Table 5). These found no significant association between Hispanic ethnicity or Spanish language and hospitalization or death when the other variable was excluded. On average, Hispanic patients are younger than the overall population, which may drive this neutral association with adverse outcomes despite higher rates of infection.

### LIMITATIONS

These findings must be interpreted in the context of the study design. The number of cases diagnosed depends heavily on testing rates, which were influenced by changing testing criteria which, in turn, were influenced by local government actions, the necessity of ensuring healthcare worker safety, prevalence of community health centers conducting outpatient testing, and awareness of existing outbreaks. For this reason, we adjusted all analyses for census tract-level per capita testing rates. Additionally, this cohort included only patients tested within one large healthcare system, leading to under-representation of neighborhoods in this geographic area which are predominantly served by other institutions, raising the possibility of selection bias. Similarly, while we fully capture test positivity among those tested and death among those hospitalized, an individual may have been tested in the MGB system but later presented to another hospital for admission, leading to incomplete capture of the hospitalization outcome. This may be especially common among those with missing information (e.g. insurance) who may have only interacted with the MGB system briefly for a SARS-CoV-2 test. While deaths which occurred in individuals admitted to other hospitals were not captured, these individuals would have been excluded from the death analysis which only included those hospitalized in our system. Although more than 500 deaths occurred in the cohort, power to detect smaller differences related to highly correlated variables may be limited. Race is a social construct influenced by many factors; the prescribed assignment of race based on country of origin and language oversimplifies the concept of race and may lead to misassignment of individuals of mixed ancestry, complex racial identity, or a minority racial identity in their country of origin. However, in a sensitivity analysis in which only those with missing race data, but not self-reported “Other,” race were reassigned (n=393 of 3,248, Supplemental Tables 6-7), the effects of individual race on infection risk persisted. Finally, census tract-level indicators are correlated with both individual-level and other census tract-level characteristics, suggesting complex interdependencies between these variables.

## CONCLUSIONS

This study highlights the important impact of sociodemographic and economic factors on risk of SARS-CoV-2 infection, hospitalization, and death and demonstrates a method for studying socioeconomic effects in large healthcare cohorts that considers their complex interplay. These findings help to explain the racial and ethnic disparities seen during the COVID-19 pandemic as part of a complex milieu of socioeconomic and structural factors which contribute to disease risk. As many countries anticipate second waves of SARS-CoV-2 infection, the findings of this study from the first wave of SARS-CoV-2 in a large urban area may help to identify individuals and communities at greatest risk during subsequent outbreaks.

## Data Availability

Due to the nature of this research, participants of this study did not agree for their data to be shared publicly, so supporting data is not available.

## AUTHOR CONTRIBUTIONS

SJC conceived of the study. SJC and CML designed the analysis which was performed by CML. CJP oversaw the analysis. CJP, MU, and DJW provided critical feedback on all analyses. SMB provided critical feedback regarding the use of race or ethnicity in the analysis and the interpretation of the results. SJC drafted the manuscript with critical review provided by all authors.

